# Grading of Erythema and Visual Attributes in Atopic Dermatitis across Diverse Skin Tones Using a Vision AI Pipeline

**DOI:** 10.64898/2026.03.30.26349755

**Authors:** Mahla Abdolahnejad, Manuella Kyremeh, James Smith, Gloria Fang, Hannah O. Chan, Rakesh Joshi, Colin Hong

**Affiliations:** Skinopathy Research, Skinopathy Inc., North York, Ontario, Canada; Department of Physiology, Faculty of Medicine, University of Toronto, Ontario, Canada; Faculty of Information Technology, Monash University, Melbourne, Victoria, Australia

## Abstract

**Background:** Atopic dermatitis (AD) is a prevalent chronic inflammatory skin disease associated with clinical, psychosocial, and economic burden. Accurate severity assessment is essential for guiding treatment escalation and monitoring disease activity, yet clinician-based scoring systems such as the Eczema Area and Severity Index (EASI) are limited by subjectivity and considerable inter- and intra-rater variability. Erythema, a key driver of AD severity grading, is particularly prone to inconsistent evaluation due to differences in ambient lighting, device quality, skin tone, and rater experience, underscoring the need for objective, reproducible assessment tools.

**Objective:** To develop and validate an artificial intelligence (AI) pipeline for grading erythema, excoriation, and lichenification severity in AD from clinical photographs. The study evaluated the level of agreement between AI severity ratings in each category against dermatologists, non-specialists, and a consensus reference standard, with erythema as the primary outcome of interest.

**Methods:** A two-stage AI pipeline was developed using EfficientNet B7 convolutional neural networks (CNNs). The first CNN was trained as a binary AD classifier on 451 AD and 601 non-AD images for lesion detection and segmentation. The second CNN was trained on 173 dermatologist-annotated AD images which were scored on a 0–3 ordinal scale for erythema, excoriation, and lichenification. This CNN had downstream, feature extraction algorithms such red channel contrast for erythema, Law’s E5L5 for excoriation, and S5L5 texture maps for lichenification. In a cross-sectional validation study, 41 independent test images were scored by two blinded dermatologists and two blinded physicians. AI predictions were compared to individual rater groups and mode-derived consensus scores using weighted Cohen’s kappa (κ), classification accuracy, confusion matrices, and error direction analyses.

**Results:** On internal validation, the severity CNN achieved 84% overall accuracy (averaged across all three attributes), 86% sensitivity, 87% specificity, and a macro-averaged area under the receiver operating characteristic curve (AUC) of 0.90. In the external comparison with blinded human raters, erythema agreement between the AI and dermatologist consensus was substantial (accuracy 80.7%; κ = 0.68), with no large (≥2-point) misclassifications. Physician consensus agreement was lower (accuracy 54.8%; κ = 0.34), reflecting greater variability among primary care physicians (non-specialists). For excoriation, AI–dermatologist agreement was moderate (accuracy 72.4%; κ = 0.62); for lichenification, agreement was similar (accuracy 71.4%; κ = 0.59). Across all features, disagreements were predominantly between adjacent severity categories. The AI was able to generate erythema severity grades for images of darker skin tones that dermatologists typically would not rate and were marked as “unable to assess.”

**Limitations:** The validation set was small (41 images), severe cases (score 3) were underrepresented, one rater participated in both training annotation and validation scoring, and sample size was insufficient for robust stratification by skin tone or body site.

**Conclusion:** The AI pipeline demonstrated dermatologist-level accuracy for erythema scoring, consistent moderate agreement for excoriation and lichenification, and a potential advantage in assessing erythema on darker skin tones. These findings support its potential as a standardized, objective tool for AD severity assessment. Prospective validation in larger, more diverse cohorts is warranted.

## Introduction

Atopic dermatitis (AD) is a chronic, relapsing inflammatory skin disease defined by barrier dysfunction and eczematous lesions whose morphology and distribution shift across the lifespan (Tian et al., 2023). Pruritus, pain, anxiety, depression, and sleep disturbance frequently accompany the cutaneous disease (Kim, J. et al., 2019; *Courtney, A., & Su, J. C., 2024*) and roughly 204 million people worldwide are affected (an estimated 15–20% of children and 2–5% of adults) with an annual economic burden exceeding $5 billion in the United States alone (Tian et al., 2023; Adamson, 2024; Faye et al., 2024). By any measure, AD is a major public health problem requiring both dependable measures for disease activity tracking and effective management.

Clinical assessment of AD rests on four cardinal signs: erythema, excoriation, lichenification, and oedema/papulation. Erythema is the most immediately visible sign and is highly correlated with active inflammation. Excoriation is a sign of scratching driven by pruritus; lichenification signals epidermal thickening from chronic rubbing; and finally, oedema/papulation reflects the underlying dermal inflammatory infiltration (Kim, J. et al., 2019). Treatment escalation, from topical management to systemic agents, phototherapy, biologic therapy, as well as follow-up frequency, and clinical trial endpoints all depend on reliable grading of these signs (Ameen et al., 2024; Waligóra-Dziwak, K., et al., 2026). If the measurement is unreliable, the clinical decision it supports is compromised.

### Limitations of Current Scoring Instruments

The Eczema Area and Severity Index (EASI) is the consensus core outcome instrument for AD clinical trials, endorsed by the Harmonising Outcome Measures for Eczema (HOME) initiative on the basis of its validity, responsiveness, and internal consistency (Hanifin, J. M. et al., 2008; Chalmers et al., 2014; Chopra et al., 2017; Hanifin, J. M. et al., 2022). EASI rates erythema, infiltration/papulation, excoriation, and lichenification on a 0–3 scale across four body regions weighted by surface area, producing a composite score from 0 to 72. The SCORing Atopic Dermatitis index (SCORAD) adds patient-reported symptoms to clinician-assessed signs, while the Investigator’s Global Assessment (IGA) offers a single-item global severity rating. Each instrument is widely used. Each also carries well-documented limitations.

The central problem is agreement. Even among trained dermatologists, concordance on individual EASI sign scores is frequently moderate at best. Bożek and Reich (2017) found acceptable intra-rater reliability for total EASI but variable inter-rater agreement on individual components; none of the three scales they examined (EASI, objective SCORAD, IGA) showed a clear advantage. Waligóra-Dziwak et al. (2026) confirmed that EASI had the highest inter-rater reliability among five scoring systems (ICC = 0.768–0.796) when used by dermatology residents, yet individual sign scores still varied substantially. In the remote setting, Van Cranenburgh et al. (2023) reported good inter-rater reliability for remote EASI (ICC = 0.77) but flagged image quality, completeness, and capture standardisation as additional error sources. Across these studies, the same factors recur: ambient lighting, device characteristics, rater calibration, and clinical experience all erode scoring consistency, particularly for erythema and excoriation, where subtle colour and textural distinctions carry clinical weight (Ameen et al., 2024; Waligóra-Dziwak, K., et al., 2026).

### Artificial Intelligence in Inflammatory Skin Disease Assessment

Deep learning and computer vision have begun to offer an alternative path. In psoriasis, image-based PASI estimation has reached rater-level performance across multiple groups, with early evidence of clinical trial-to-real-world translation (Schaap et al., 2022; Huang et al., 2023; Xing et al., 2024). In acne, deep learning models trained against the IGA scale have achieved strong agreement with expert clinicians (Lim et al., 2019; Watanabe et al., 2024). These adjacent domains supply methodological precedents such as per-sign confusion analyses, interpretable feature extraction, attention to training dataset diversity, that are directly transferable to AD pipeline design.

Within AD itself, the evidence base is growing but uneven. Bang et al. (2021) showed that CNNs could score individual EASI components from approximately 8,000 standardised clinical photographs, achieving high accuracies (erythema 90.6%, excoriation 87.5%, lichenification 85.9%). But their dataset consisted entirely of tightly framed images from a single East Asian population (Fitzpatrick types II–IV) captured under controlled lighting, conditions that minimise variability and inflate performance relative to heterogeneous real-world imaging. Medela et al. (2022) proposed an automatic SCORAD system (ASCORAD) demonstrating feasibility of image-based estimation that tracked expert assessment. Maulana et al. (2024) reported 89.8% accuracy using ResNet50 on an Indonesian cohort, though this again raised questions about generalisability beyond the training population. Muhaba et al. (2021) tackled eczema-area detection as a prerequisite for automated EASI-like computation. Most recently, Okata-Karigane et al. (2025) developed an AI model for smartphone-uploaded AD images achieving 98% accuracy in body-part detection and demonstrated correlation with clinician-scored severity measures (R = 0.73, p < 0.001), supporting the feasibility of AI as a digital biomarker for real-world monitoring.

Two recent reviews crystallise what this body of work collectively shows and where it falls short. Cao et al. (2025) and Liu et al. (2025) both highlight that AD-AI studies are predominantly retrospective, often single-centre, and underpowered for subgroup analysis by skin tone or body site. External validation with standardised capture protocols reflecting real clinical variation remains scarce. Meta-analytic evidence from dermatology AI broadly confirms that model performance drops when moving from benchmark to practice settings (Nadour et al., 2025). The gap between laboratory accuracy and clinical utility persists.

### Erythema Detection in Skin of Colour

Erythema detection poses a particular clinical challenge across the skin tone spectrum. In darker skin, inflammation often does not present as bright red. Instead, it may appear as hyperpigmented violet, grey, or reddish-brown relative to surrounding unaffected skin (Adawi et al., 2023;– Chiricozzi, A. et al., 2023). Hence, even experienced dermatologists can underrecognize erythema in richly pigmented skin leading to the systematic underestimation of AD severity and delayed treatment escalation (Forsyth et al., 2025; Wilson et al., 2022; Kaufman et al., 2018). Chaudhary and Agrawal (2024) showed that the disparity in AD diagnosis between Black and White populations is partly attributable to erythema being concealed on darker skin tones, contributing to later and more severe disease presentation. Servattalab et al. (2024) demonstrated that non-Hispanic Black and Hispanic infants with AD experience significant disparities in time to dermatologist referral and care access compared with non-Hispanic White infants.

The instruments themselves may compound the problem. EASI and SCORAD were largely developed and validated using images featuring lighter skin tones and may not capture inflammatory chromatic changes in skin of colour (Kamp et al., 2025; Bissonnette et al., 2023). Forsyth et al. (2025) have argued that conventional descriptors of erythema are unreliable in Fitzpatrick phototypes IV–VI, advocating for augmented definitions and calibrated colorimetry. In parallel, most AI training datasets underrepresent Monk skin tone scale categories 7–10, which means automated erythema detectors risk inheriting or amplifying existing diagnostic biases unless explicitly evaluated across the full tone spectrum (Daneshjou et al., 2022).

Paediatric AD introduces additional complexity where lesion morphology and distribution in infants and young children differ from adult patterns. Pediatric AD often has a predilection for the face, scalp, and extensor surfaces in early childhood that transitions to classic flexural involvement later (Yew et al., 2019). The subtler and more diffuse presentation of erythema in paediatric patients complicates visual grading, and the scarcity of paediatric-specific training images further limits generalisable AI development for this population.

### Study Objective

To address gaps directly a validated AI pipeline that derives quantitative severity measurements of erythema, excoriation, and lichenification from heterogeneous, web-sourced clinical photographs and compared them against multi-rater human scoring was developed. This pipeline employs an EfficientNet B7 architecture for both AD detection and severity grading, combined with interpretable, attribute-specific feature extraction algorithms (red-channel contrast for erythema; Law’s texture energy maps for excoriation and lichenification). Unlike prior AD-AI studies that relied on tightly controlled, single-population imaging conditions, this study uses images spanning diverse skin tones, anatomical sites, and capture environments, and benchmarks performance against four independent blinded raters, two dermatologists and two primary-care physicians. Erythema is designated as the primary outcome given its clinical primacy, its role as a key driver of EASI scoring, and the documented challenges in its assessment across skin tones. The aim is to determine whether AI-based erythema grading can approximate dermatologist assessment and serve as a complementary tool for more objective and equitable severity evaluation in atopic dermatitis.

## Methods

### Study Design

This was a cross-sectional study comprising of two components: (1) the development and internal validation of a multi-stage AI pipeline for scoring AD severity features from clinical photographs, and (2) an external comparison of AI-derived scores against independent human rater assessments, with erythema as the primary outcome, and excoriation and lichenification as secondary outcomes.

### Ethics and Data Governance

This was an investigator-led study using publicly available, de-identified clinical photographs of AD lesions sourced from open-access repositories. As no identifiable patient data were collected or utilised and all images were already in the public domain, formal institutional review board (IRB) approval was not required. Faces and other potentially identifying features were cropped or excluded during image curation to ensure participant anonymity.

### AI Algorithm Architecture

The AI pipeline consisted of three sequential stages: (1) AD lesion detection and segmentation, (2) severity feature learning, and (3) attribute-specific feature extraction and score derivation (Figure 1). Both neural network components employed the EfficientNet B7 architecture.

**Figure 1.**
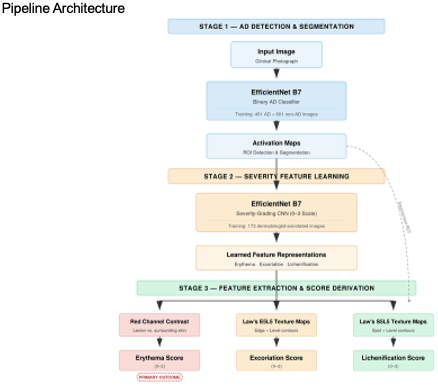

#### Stage 1: AD Detection and Lesion Segmentation

An EfficientNet B7 convolutional neural network (CNN) was trained as a binary classifier to distinguish AD lesions from non-AD skin. The training dataset comprised 451 AD images and 601 non-AD images where the non-AD category included both normal skin and images of AD-like conditions to improve discriminative specificity. This network generated activation maps used for skin segmentation and region-of-interest (ROI) detection, defining the spatial boundaries of AD-affected skin within each photograph for downstream severity grading.

#### Stage 2: Severity-Grading CNN

A second EfficientNet B7 CNN was trained on a subset of the AD image corpus that had been independently annotated by dermatologists for three visual attributes: erythema, excoriation, and lichenification. Each attribute was on a 0–3 ordinal severity scale (0 = none, 1 = mild, 2 = moderate, 3 = severe). The annotated training dataset comprised 173 images (Figure 2). The severity distribution for erythema was: score 0, n = 6; score 1, n = 61; score 2, n = 70; score 3, n = 36. For excoriation: score 0, n = 59; score 1, n = 51; score 2, n = 37; score 3, n = 26. For lichenification: score 0, n = 65; score 1, n = 51; score 2, n = 41; score 3, n = 16. These distributions are summarised in Table 1.

**Figure 2.**
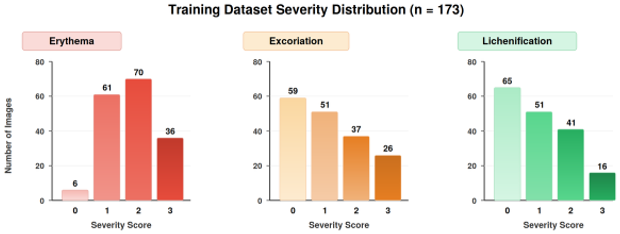

**Table 1.** Per-Class Erythema Performance Metrics (AI vs Dermatologist Consensus, n=31)

| Class | Precision | Recall | F1-Score | n |
| --- | --- | --- | --- | --- |
| 0 (None) | 1.00 | 1.00 | 1.00 | 1 |
| 1 (Mild) | 1.00 | 0.89 | 0.94 | 9 |
| 2 (Moderate) | 0.88 | 0.78 | 0.82 | 18 |
| 3 (Severe) | 0.33 | 0.67 | 0.44 | 3 |
| Accuracy | — | — | 0.81 | 31 |
| Macro avg | 0.80 | 0.83 | 0.80 | 31 |
| Weighted avg | 0.86 | 0.81 | 0.83 | 31 |

#### Stage 3: Feature Extraction and Score Derivation

Following lesion segmentation (Stage 1) and severity feature learning (Stage 2), the pipeline applied three parallel, attribute-specific algorithms to the detected ROI to derive final severity scores (Figure 1):

Erythema: Contrast in the red channel of the segmented lesion region was computed relative to surrounding unaffected skin. This quantitative redness metric, informed by the learned representations of the severity CNN, was mapped to the 0–3 erythema severity scale.

Excoriation: Contour features were extracted using Law’s E5L5 (edge × level) texture energy maps applied to the segmented ROI. These texture descriptors captured the linear scratch patterns and surface disruption characteristic of excoriation and were mapped to the 0–3 severity scale.

Lichenification: Contour features were extracted using Law’s S5L5 (spot × level) texture energy maps, which captured the coarsened, thickened skin surface patterns indicative of lichenification, and were similarly mapped to the 0–3 severity scale.

An additional area score module accepted user-provided eczema area information and calculated the percentage of body surface area affected; however, this component was not the focus of the present validation study.

#### Internal Validation of the Severity CNN

The severity CNN was evaluated on an internal held-out test set of 31 images across all three attributes (erythema, excoriation, and lichenification averaged; hereafter referred to as the EEL average). Performance metrics included per-class precision, recall, and F1-score, overall classification accuracy, and multiclass receiver operating characteristic (ROC) analysis with area under the curve (AUC) computed per class as micro- and macro-averages (Figure 3). The EEL-averaged results were: overall accuracy 84%, sensitivity (macro recall) 86%, specificity 87%, micro-average AUC 0.89, and macro-average AUC 0.90. Per-class performance was as follows: class 0—precision 1.00, recall 1.00, F1 1.00 (support n = 1); class 1—precision 0.88, recall 0.88, F1 0.88 (support n = 8); class 2—precision 0.81, recall 0.87, F1 0.84 (support n = 15); class 3—precision 0.83, recall 0.71, F1 0.77 (support n = 7). Per-class AUC values were: class 0 = 1.00, class 1 = 0.92, class 2 = 0.84, class 3 = 0.84.

**Figure 3.**
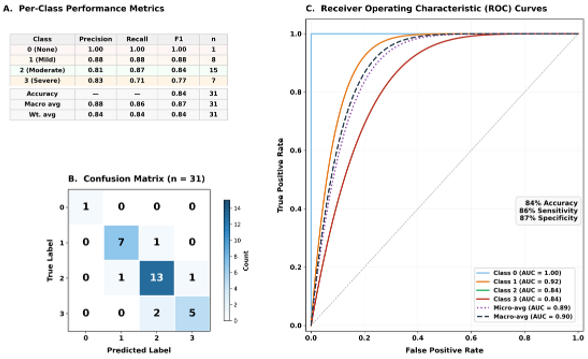

### External Validation: Human Rater Comparison

#### Rater Panel and Scoring Protocol

A set of 41 AD images, independent of the training and internal validation sets, was assembled for external validation. Each image was independently scored by four blinded clinicians: two board-certified dermatologists and two primary care physicians (non-dermatology). Raters were blinded to the AI predictions and to each other’s scores throughout the rating process. Each rater scored every image for erythema, excoriation, and lichenification on the same 0–3 ordinal scale used for algorithm training. Mode-derived consensus scores were computed separately for dermatologists, primary care physicians, and the overall panel (all four raters), and served as the reference standards for AI comparison.

### Statistical Analysis

Agreement between AI predictions and each consensus reference (dermatologist, physician, overall) was assessed using weighted Cohen’s kappa (κ) with linear weighting, overall classification accuracy (proportion of exact matches), and per-class precision, recall, and F1-score. Weighted Cohen’s kappa served as the primary measure of inter-rater agreement throughout all analyses. Confusion matrices were generated for each pairwise comparison. Error direction analysis classified each disagreement as an over-prediction or under-prediction relative to the consensus, and large errors were defined as discrepancies of ≥2 ordinal categories. Inter-rater agreement between dermatologists was separately quantified using weighted κ to benchmark human variability. Erythema was the designated primary outcome and excoriation and lichenification were secondary outcomes. Exploratory subgroup analyses by skin tone, body site, and severity stratums were planned but were limited by sample size and are reported descriptively where feasible. All analyses were performed in Python using scikit-learn.

### Erythema Assessment in Darker Skin Tones

A subset of validation images depicting AD lesions on darker skin tones (Monk scale categories 7–10) was examined as an exploratory analysis. Dermatologist assessments and AI-generated erythema scores were compared qualitatively, with particular attention to cases in which human raters were unable to assign a severity grade. The AI pipeline’s erythema maps (heatmap overlays derived from the red-channel contrast algorithm) were visually inspected to assess whether the model could detect inflammatory chromatic changes not readily apparent to clinicians on standard clinical photographs.

## Results

### Internal Validation of the Severity CNN

On the internal held-out test set (n = 31 images), the severity CNN achieved an overall classification accuracy of 84% when averaged across all three attributes (erythema, excoriation, and lichenification; EEL average). Macro-averaged sensitivity was 86%, specificity was 87%, and the macro-averaged area under the receiver operating characteristic curve (AUC) was 0.90 (micro-average AUC = 0.89). Per-class performance was highest for score 0 (precision 1.00, recall 1.00, F1 1.00; n = 1) and score 1 (precision 0.88, recall 0.88, F1 0.88; n = 8), and remained strong for score 2 (precision 0.81, recall 0.87, F1 0.84; n = 15). Performance was lowest for score 3 (precision 0.83, recall 0.71, F1 0.77; n = 7), consistent with the smaller representation of severe cases in the training corpus. Per-class AUC values were 1.00 (class 0), 0.92 (class 1), 0.84 (class 2), and 0.84 (class 3) (Figure 3).

### External Validation: AI Versus Human Rater Comparisons

A total of 41 images were presented to the rater panel. After exclusion of images that one or more raters were unable to score, the analysable sample sizes were 31 images for erythema, 29 for excoriation, and 28for lichenification. Across all three attributes, a score of 2 (moderate severity) was the most frequently assigned category by all rater groups, including the AI. Primary Care Physicians were more likely than either the AI or dermatologists to assign a score of 3 (severe), particularly for excoriation and lichenification, a pattern that contributed to the lower AI–physician agreement observed across analyses. Scores of 0 (none) were assigned infrequently by all raters across all attributes.

### Erythema (Primary Outcome)

The severity score distributions for erythema are presented in Figure 4A. All rater groups, including the AI, most frequently assigned a score of 2 (moderate erythema). Physicians assigned a score of 3 (severe erythema) more often than either the AI or dermatologists.

**Figure 4.**
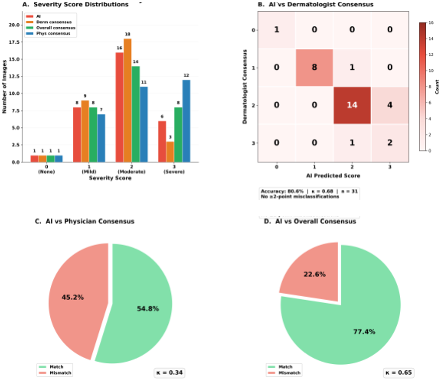

Agreement between AI predictions and dermatologist consensus was substantial. The AI achieved an exact match rate of 80.6% (25 of 31 images), with a weighted Cohen’s κ of 0.68 (Figure 4B; Table 2A). Per-class metrics (Table 1) showed high precision and recall for scores 0 (precision 1.00, recall 1.00; n = 1), 1 (precision 1.00, recall 0.89; n = 9), and 2 (precision 0.88, recall 0.78; n = 18). Performance was weakest for score 3 (precision 0.33, recall 0.67; n = 3), reflecting the small number of severe cases in the validation set. The overall accuracy was 0.81, with a macro-averaged precision of 0.80, macro-averaged recall of 0.83, weighted-average precision of 0.86, and weighted-average recall of 0.81. Agreement was strongest for moderate erythema (score 2), where 14 of 18 cases were correctly classified. Misclassifications were confined to adjacent categories; the most common error was moderate cases predicted as severe.

**Table 2.**
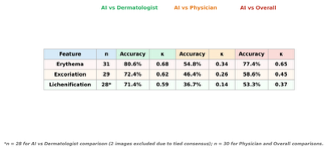
Cross-Feature Agreement Summary AI vs Human Rater Consensus.

Agreement between AI predictions and primary care physician consensus was lower, with an exact match rate of 54.8% and κ = 0.34 (Figure 4C). These physicians were more likely than the AI to assign both higher and lower severity scores, indicating greater variability in primary care physician assessment.

Against the overall consensus (all four raters), the AI achieved an exact match rate of 77.4% and κ = 0.65 (Figure 4D).

Error analysis revealed no large (≥2-point) discrepancies between AI predictions and dermatologist consensus across the 31 erythema images (Table 3). The mean difference between AI and consensus scores was −0.03, with a median difference of 0.0. Of the 31 images, 77.4% were exact matches, 12.9% were under-predicted by one category, and 9.7% were over-predicted by one category. Among the seven disagreements, the AI was more likely to under-predict (4 of 7, 57.1%) than to over-predict (3 of 7, 42.9%), though this difference was not statistically significant given the small sample.

**Table 3.** Error Direction and Magnitude Analysis (AI vs Overall Consensus)

| Feature | n | ≥2-pt Errors | % Large Errors | Mean Diff | Median Diff | Exact Match % | Under-predicted | Over-predicted |
| --- | --- | --- | --- | --- | --- | --- | --- | --- |
| Erythema | 31 | 0 | 0% | -0.03 | 0.0 | 77.4% | 12.9% | 9.7% |
| Excoriation | 29 | 2 | 8.3% | -0.25 | 0.0 | 58.6% | — | — |
| Lichenification | 30 | 2 | 7.7% | +0.15 | 0.0 | 53.3% | — | — |

### Excoriation (Secondary Outcome)

For excoriation, the analysable sample comprised 29 images after exclusion of cases that raters were unable to score. Severity score distributions (Figure 5A) showed that the AI favoured score 1 (mild) more frequently than the consensus groups, whereas physicians and overall consensus tended toward scores 2 and 3.

**Figure 5.**
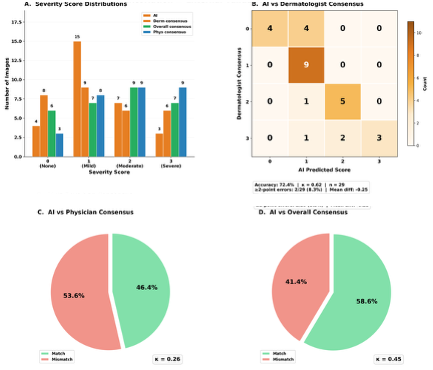

Agreement between AI and dermatologist consensus was moderate, with an exact match rate of 72.4% and κ = 0.62 (Figure 5B; Table 2B). Against primary care physician consensus, agreement was substantially lower (accuracy 46.4%, κ = 0.26; Figure 5C), and against the overall consensus the AI achieved 58.6% accuracy with κ = 0.45 (Figure 5D).

Error analysis identified 2 large (≥2-point) discrepancies out of 29 images (8.3%). The mean difference between AI and overall consensus scores was −0.25, with a median of 0.0 (Table 3).

### Lichenification (Secondary Outcome)

For lichenification, 30 images were analysable; two additional images were excluded from the AI–dermatologist comparison due to tied dermatologist consensus, yielding 28 images for that analysis. Score distributions (Figure 6A) showed that both the AI and dermatologist consensus most frequently assigned scores of 1 and 2, whereas primary care physicians more often assigned score 3.

**Figure 6.**
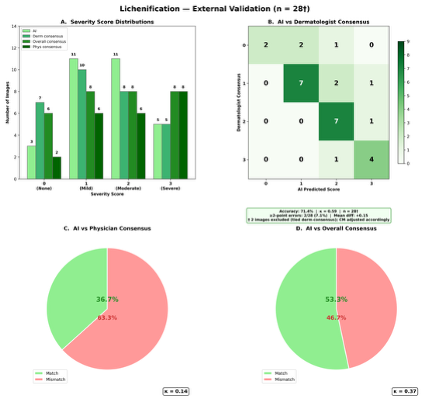

AI–dermatologist consensus agreement was moderate, with an exact match rate of 71.4% and κ = 0.59 (Figure 6B; Table 2C). Against physician consensus, agreement was weak (accuracy 36.7%, κ = 0.14; Figure 6C), and against the overall consensus the AI achieved 53.3% accuracy with κ = 0.37 (Figure 6D).

Two large (≥2-point) discrepancies were observed out of 30 images (7.7%). The mean difference between AI and overall consensus scores was +0.15, with a median of 0.0 (Table 3).

### Cross-Feature Comparison

Table 2 summarises agreement metrics across all three clinical features. Erythema consistently yielded the highest AI–human agreement irrespective of the reference standard used, followed by excoriation and then lichenification. Across all features, AI alignment was substantially stronger with dermatologist consensus than with physician consensus. This pattern held for both accuracy and weighted κ.

Error severity was generally contained across all features (Table 3). For erythema, no large (≥2-point) discrepancies were observed; for excoriation and lichenification, large errors occurred in 8.3% and 7.7% of images, respectively (2 images each). The mean score difference between AI and consensus was close to zero for erythema (−0.03) and lichenification (+0.15) but showed a modest negative shift for excoriation (−0.25).

### Erythema Assessment in Darker Skin Tones

An exploratory qualitative comparison of AI and dermatologist erythema scoring was conducted on a subset of validation images depicting AD lesions on darker skin tones (Figure 7). Three illustrative cases are presented. In Case A, depicting a lesion on moderately pigmented skin with visible erythema, both the dermatologist and the AI assigned a severity grade of 3 (severe); the AI-generated erythema heatmap showed strong activation over the affected regions, consistent with the clinical assessment. In Cases B and C, depicting lesions on deeply pigmented skin, dermatologists marked erythema as “unable to assess.” The AI pipeline generated erythema heatmaps for both images and assigned a severity grade of 1 (mild) to each, detecting subtle chromatic changes in the red channel that were not apparent to the clinicians on visual inspection of the standard photographs.

**Figure 7.**
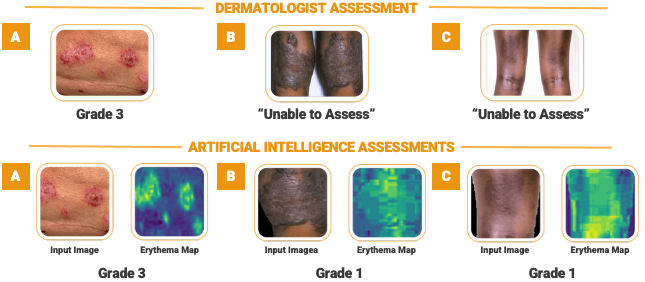

These cases suggest that the AI’s red-channel contrast algorithm can detect low-level inflammatory colour shifts in darker skin tones where human raters cannot render a clinical judgment. The sample was too small for quantitative inference, but the observation supports the hypothesis that algorithmically derived erythema scores may complement clinician assessment in populations where conventional visual evaluation is limited by skin pigmentation.

## Discussion

The principal finding of this study is that an EfficientNet B7–based AI pipeline achieved substantial agreement with dermatologist consensus for erythema grading in atopic dermatitis, with an exact match rate of 80.7% and a weighted Cohen’s κ of 0.68 across 31 analysable images. No large misclassifications (≥2 ordinal categories) were observed. The mean score difference was −0.03 (median 0.0), with under-predictions (12.9%) and over-predictions (9.7%) confined to adjacent categories – indicating no systematic directional bias. Agreement with physician consensus was substantially lower (accuracy 54.8%; κ = 0.34), and agreement with the combined four-rater consensus was 77.4% (κ = 0.65). In short, the algorithm tracked dermatologists more closely than it tracked primary care physicians, which is precisely the behaviour a clinically useful tool should exhibit.

Performance on secondary features was more modest, and this is expected. AI–dermatologist agreement for excoriation was 72.4% (κ = 0.62) and for lichenification 71.4% (κ = 0.59); physician consensus agreement dropped to κ = 0.26 and 0.14, respectively. A small number of large errors were observed for both features (2 of 29 for excoriation; 2 of 28 for lichenification). These patterns do not point to a failure unique to the algorithm. They reflect the inherent difficulty of standardising features that are more subjective even among trained human raters. On internal validation, the severity CNN achieved 84% accuracy, 86% sensitivity, 87% specificity, and a macro-averaged AUC of 0.90, though per-class performance was weakest for score 3 (precision 0.33, recall 0.67 for erythema in the external comparison), a consequence of limited severe-case representation.

### Comparison with Existing Literature

The erythema accuracy of 80.7% and κ of 0.68 are lower than figures reported by Bang et al. (2021), who achieved 90.6% for erythema using a CNN trained on approximately 8,000 photographs from a single East Asian population, and by Maulana et al. (2024), who reported 89.8% using ResNet50 on an Indonesian cohort. However, both studies used tightly controlled, single-population imaging conditions that suppress variability. The pipeline in this study was evaluated on heterogeneous, web-sourced and real patient images spanning diverse skin tones, anatomical sites, and capture environments. This design choice traded absolute accuracy for demographic diversity and is not a limitation. It is a deliberate decision grounded in the well-documented gap between benchmark performance and clinical deployment. Meta-analytic evidence from Nadour et al. (2025) confirms that dermatology AI performance decreases when moving from curated datasets to practice settings, and the present results are consistent with that pattern.

The moderate agreement for excoriation (κ = 0.62) and lichenification (κ = 0.59) mirrors published inter-rater reliability data for these features among human clinicians. Bożek and Reich (2017) reported variable inter-rater agreement on individual EASI components, and Waligóra-Dziwak et al. (2026) confirmed that while total EASI inter-rater reliability was acceptable (ICC = 0.768–0.796), individual sign scores showed substantial scatter. The lower AI–human agreement for excoriation and lichenification therefore reflects measurement difficulty inherent to these features rather than isolated algorithmic failure.

Architecturally, the pipeline differs from most prior AD-AI systems in a way that matters for clinical adoption. It combines EfficientNet B7 feature learning with interpretable, attribute-specific extraction algorithms: red-channel contrast for erythema and Law’s texture energy maps for excoriation (E5L5) and lichenification (S5L5). This design enables clinicians to inspect what the model is detecting and why, thus, it provides a degree of transparency that black-box systems cannot offer. Recent reviews have emphasised interpretability as a prerequisite for clinical trust (Liu et al., 2025; Cao et al., 2025). The use of four independent blinded raters as the reference standard strengthens the clinical relevance of these comparisons relative to studies relying on a single expert.

One exploratory finding warrants particular attention. The AI generated erythema scores for images on darker skin tones (Monk 7–10) where dermatologists marked erythema as “unable to assess.” The red-channel contrast algorithm detected subtle chromatic shifts not visible on standard clinical inspection. This observation aligns with evidence that erythema is systematically underestimated in darker phototypes (Forsyth et al., 2025; Wilson et al., 2022; Chaudhary and Agrawal, 2024) and that EASI and SCORAD were largely validated on lighter skin tones (Kamp et al., 2025; Bissonnette et al., 2023). If this capability is confirmed in adequately powered prospective studies, it could address a significant and well-documented equity gap in AD assessment. Importantly, the “unable to assess” designation reflects the constraints of the scoring protocol rather than a deficiency in the physician’s clinical judgment; dermatologists who omit erythema from their rating may still form a global impression of disease severity that incorporates cues beyond the single sign. Future studies should therefore compare the physician’s overall severity rating for these images against the AI’s sign-level scores to determine whether the model’s detection of mild erythema aligns with or falls short of the holistic clinical assessment. Even where the AI detects erythema, it may still under-estimate severity relative to the physician’s global inference, which directly impacts the accuracy and clinical utility of the model in darker skin tones.

### Clinical Implications

The practical question is where this pipeline fits. The substantial erythema agreement with dermatologist consensus, combined with the absence of large misclassifications, positions it as a decision-support adjunct in settings where dermatologist access is limited. In tele-dermatology and remote monitoring, where Van Cranenburgh et al. (2023) demonstrated acceptable but imperfect inter-rater reliability for remote EASI (ICC = 0.77), a standardised algorithmic tool could reduce measurement variability and support longitudinal tracking. Integration into an EASI-based framework that combines AI-derived sign scores with body-surface-area information would enable automated composite severity scores directly comparable to established trial endpoints. For patient-facing applications, the feasibility of smartphone-based severity estimation demonstrated by Okata-Karigane et al. (2025) suggests such pipelines could support more frequent monitoring and earlier flare identification for earlier intervention.

The algorithm should not be positioned as a replacement for clinical judgment. The weaker performance for severe erythema (score 3) and the modest agreement for excoriation and lichenification make this clear. Human oversight remains essential, particularly at severity extremes and when treatment decisions hinge on the result. A human-in-the-loop deployment model, in which AI-generated scores inform but do not determine treatment decisions, would be consistent with current regulatory frameworks for clinical decision-support software and with the evidence presented here.

### Limitations

Images that raters could not score were excluded, which may have introduced optimistic selection bias. Although images spanned the Monk skin tone scale, sample size was insufficient for reliable performance stratification by tone, a gap that has been repeatedly identified in the field (Daneshjou et al., 2022; Kamp et al., 2025). Moreover, web-sourced images introduced uncontrolled variability in resolution, lighting, and capture conditions. Another consequence of the reduced size of the validation dataset is limited statistical power (41 images; 31, 29, and 28 analysed for erythema, excoriation, and lichenification, respectively). Furthermore, severe cases (score 3) were underrepresented in both training and validation sets, thus the most clinically consequential disease states were precisely those for which confidence was lowest. Similarly, mild presentations (score 0 and 1) were relatively scarce for certain attributes, introducing a potential bias toward moderate severity in the training distribution; future datasets should include a broader spectrum of mild cases, which could be obtained through crowdsourcing or primary care recruitment. One rater participated in both training annotation and validation scoring, introducing potential circularity bias that may have inflated performance estimates. Overall, the cross-sectional design prevents assessment of responsiveness to longitudinal change or treatment effect in this study.

### Future Directions

Several priorities follow from these findings. First, larger and more diverse datasets are needed, with greater representation of severe cases, darker skin tones (Monk 7–10), paediatric patients, and varied anatomical sites and capture devices. Increasing the number of independent raters and enforcing strict separation between training annotators and validation raters will strengthen reference standards. Second, integration into an EASI-based scoring framework combining AI-derived sign severity with structured body-surface-area information would produce composite scores directly comparable to trial endpoints. Third, the exploratory observations on darker skin tone assessment require formal evaluation in adequately powered prospective cohorts. Fourth, prospective, multi-centre longitudinal validation is essential to assess whether the pipeline can detect treatment-induced change across real-world clinical environments, including primary care, tele-dermatology, and patient self-monitoring. Fifth, implementation studies assessing clinician and patient acceptance, workflow integration, and the impact of AI outputs on treatment decisions will be critical for translation into routine clinical use. Sixth, future work should compare AI-derived sign-level severity scores against global dermatologist severity ratings or weighted composite scores to determine whether sign-level accuracy translates into meaningful differences at the global severity scale, which directly impacts treatment stratification and clinical trial eligibility criteria. Seventh, the training dataset should be expanded to include a greater proportion of mild presentations (scores 0 and 1), as the current distribution is biased toward moderate severity; recruitment from primary care settings or crowdsourced image collection may be effective strategies for addressing this gap.

### Conclusion

This study demonstrates that an Vision–based AI pipeline can achieve substantial agreement with dermatologist consensus for erythema grading in atopic dermatitis, with an exact match rate of 80.7% and a weighted Cohen’s κ of 0.68, and no large misclassifications. Agreement for excoriation and lichenification was moderate, consistent with the inherent subjectivity of these features even among trained human raters. The AI’s ability to generate erythema severity grades for images on darker skin tones where dermatologists were unable to render a clinical judgment highlights its potential to address documented equity gaps in AD assessment. However, the small validation sample, underrepresentation of both severe and mild cases, and cross-sectional design limit the generalisability of these findings. Prospective validation in larger, more diverse cohorts, with expanded representation of mild presentations, darker skin tones, and paediatric patients, is essential. Comparison of AI-derived sign-level scores against global dermatologist severity ratings will be critical to determine whether sign-level accuracy translates into meaningful clinical impact on treatment stratification and trial eligibility. With further validation and integration into EASI-based scoring frameworks, this pipeline has the potential to serve as a standardised, objective decision-support tool for AD severity assessment across diverse clinical settings.

## Data Availability

All data produced in the present work are contained in the manuscript; All other data, including non-open sources images, as these are PHI, are disallowed by the REBs, unless we have explicit patient consent.

